# Point-of-care semiquantitative test for adherence to tenofovir alafenamide or tenofovir disoproxil fumarate

**DOI:** 10.1101/2021.08.16.21262133

**Authors:** Derin Sevenler, Xin Niu, Sandy Dossantos, Mehmet Toner, Tim R. Cressey, Rebecca Sandlin, Paul Drain

**Author notes:** Some elements of this work have been presented publicly at the Conference on Retroviruses and Opportunistic Infections (virtual, March 6-10, 2021) and the Consortium of Universities for Global Health (virtual, March 12-14, 2021).

## Abstract

**Objective:** Objective measurement of antiretrovirals may aid clinical interventions for improving adherence to HIV prevention or treatment regimens. A point-of-care urine test could provide real-time information about recent adherence to regimens containing tenofovir disoproxil fumarate (TDF) or tenofovir alafenamide (TAF). We developed a lateral flow immunoassay (LFA) and enzyme-linked immunoassay (ELISA) for urinary tenofovir.

**Methods:** Intensity of the LFA test line was quantified using an optical reader and visually scored 0 – 5 by two independent people, using a reference card. The sensitivity and specificity of both the ELISA and LFA were determined for two different tenofovir concentration cutoffs for TDF and TAF adherence—1,500 ng/mL and 150 ng/mL, respectively. To validate the assays, we measured 586 urine samples from 28 individuals collected as part of a study of tenofovir pharmacokinetics in adults, which were also measured by mass spectrometry as ground truth.

**Results:** Both the LFA signal and ELISA signal were each strongly correlated to drug concentrations (0.91 and 0.92 respectively). The LFA signal and ELISA were highly sensitive and specific at both thresholds (LFA se/sp TDF 89%/96%, TAF 90%/96%; ELISA se/sp TDF 94%/94%, TAF 92%/84%). Visual scoring of the LFA was also highly sensitive and specific at both the TDF and TAF thresholds (se/sp TDF 91%/94%, TAF 87%/90%).

**Conclusions:** Our rapid semi-quantitative test can measure TFV concentrations relevant to both TAF or TDF adherence, which may support adherence-promoting interventions across a range of HIV care settings.

## Introduction

Oral antiretroviral therapy (ART) is highly effective for people living with HIV (PLWH) to suppress viral replication and prevent onward transmission, but monitoring and improving drug adherence have been challenging. The efficacy of ART in suppressing HIV RNA viral load is highly dependent on maintaining therapeutic concentrations of the drug, and assaying drug concentrations may also help to estimate current disease burden or identify risk of future viremia.^1,2^ As compared to subjective measures of adherence, such as self-reported adherence, clinician assessments, pill counts, pharmacy refill records, and clinic visit attendance, direct measurement of drug concentrations is the most accurate and objective method for assessing uptake of oral ART and PrEP.^4,5^ Therefore, accurate real-time information may empower effective targeted interventions to improve adherence.^6^

The drug tenofovir (TFV) is frequently included in first-line oral ART and PrEP regimens, formulated as one of two prodrugs, tenofovir disoproxil fumarate (TDF) or tenofovir alafenamide (TAF).^7^ TFV in the urine is highly correlated with days since the last dose of TDF, with a washout half-life of about 17 hours.^8^ Rapid lateral flow tests for TFV in urine have recently been reported.^9–11^ These tests accurately provide a binary ‘yes/no’ visual readout of TFV concentrations above or below the TDF threshold concentration of 1,500 ng/mL. However, a test which can discriminate lower TFV concentrations in the urine would be able to differentiate between moderate and very low adherence to TDF, and furthermore may be valuable for determining adherence to TAF.^12^

## Objective

We sought to develop a lateral flow assay for TFV in urine, and evaluated two different readout modalities to enable a semiquantitative test relevant to both TDF and TAF adherence. It is reported elsewhere that about 98% of individuals with a TFV concentration in the urine of less than 1,500 ng/mL have not taken a dose in over 24 hours, and about half of these individuals have not taken a dose in over 3 days.^13^ In the case of TAF, the half-life of TFV in the urine is about 30 hours and likewise highly correlated with adherence since the biophysical mechanisms of TFV clearance are similar as in TDF.^14^ Urine concentrations of TFV are roughly 10-fold lower with recommended dosages of TAF (10-25 mg, once daily) compared to standard doses of TDF (300 mg, once daily).^15,16^ Therefore, we chose a threshold concentration of 150 ng/mL of urine TFV for TAF adherence specifically.

## Patients and Methods

### Study Population

The TARGET Study was a randomized, open-label, clinical pharmacokinetic study of tenofovir in healthy adult volunteers without HIV or Hepatitis B infection in Thailand.^17^ Urine samples were collected from 28 participants taking oral TDF (300 mg) and emtricitabine (200 mg) randomly assigned to one of three regimens: once daily, four times weekly, or 2 times weekly. Urine samples were collected during the six-week treatment phase as well as a subsequent 4-week washout phase.

### Laboratory Procedures

#### Tenofovir quantification

TFV concentration in each urine sample was measured previously by liquid chromatography-tandem mass spectrometry (LC-MS/MS).^18^ TFV urine concentrations were below the LC-MS/MS lower limit of quantification (LLOQ) of 50 ng/mL in 79 samples, and above the Upper Limit of Quantification (ULOQ) of 23,200 ng/mL in 21 samples. Deidentified sample aliquots were stored frozen at -80°C.

#### Immunoassay development

We previously identified antibodies which bind TFV with high specific affinity.^19^ We developed and optimized a competitive enzyme-linked immunosorbent assay (ELISA) as well as competitive lateral flow immunoassay (LFA) for TFV in urine. The LC-MS/MS TFV concentrations of 74 representative urine samples were unblinded during development to guide optimization of the ELISA and LFA assay reagents and parameters, and were not included in validation studies.

#### Assay validation

Urine samples were thawed overnight at room temperature immediately prior to testing. LFA strips were read at test completion (i.e., after 10 minutes) using a commercial strip reader (Axxin Ax-2x-S), which quantified peak and overall intensities of both the test line and control line. Test line brightness was then assessed visually by two people. Visual assessments were performed independently using the same visual intensity reference card which had a graduated scale from 0 (invisible) to 5 (very dark).

### Statistical Analysis

For each ELISA microwell plate, the limit of detection (LOD) and LLOQ were determined as 3x and 6x the standard deviation of the blank sample, respectively. The ELISA standard curve for each plate was fitted with a 3-parameter logistic regression and used to interpolate TFV concentrations in those samples.

The Spearman correlation coefficient was determined between the TFV concentration as measured by LC-MS/MS, and the concentration measured by ELISA, as well as with LFA test line optical intensity.To determine the performance of the LFA as a semiquantitative test, the average test line visual score and optical reader intensity were both evaluated at two different clinically relevant threshold concentrations: 1,500 ng/mL (relevant to TDF adherence), and 150 ng/mL (relevant to TAF adherence). For both readout methods at each threshold concentration, the signal cutoff to maximize accuracy (i.e., percentage of all samples which were correctly classified at that threshold) was determined. We calculated sensitivity (percentage of samples with TFV below threshold classified correctly) and specificity (percentage of samples above threshold classified correctly) of each method at these two thresholds. We also calculated sensitivity and specificity on subsets of data excluding samples with a TFV concentration within 15% of the threshold, as measured by LC-MS/MS. These corrected values are more robust to the effect of variability in the LC-MS/MS ground-truth measurement, which had a nominal coefficient of variation of 15%.

## Results

For the ELISA, the LOD, LLOQ, IC_50_, and ULOQ were found to be 135 ng/mL, 1,055 ng/mL, 1,925 ng/mL, and 23,200 ng/mL, respectively. The Spearman correlation coefficients between ELISA *vs* LC-MS/MS was 0.92 (**Figure 1A**). For the rapid test, Spearman correlation coefficient between the LFA test line optical intensity and TFV was found to be -0.91 (**Figure 1B**; the negative correlation is an expected characteristic of competitive immunoassay). For the optical readout, the optimal threshold intensity for differentiating TFV levels above or below 1,500 ng/mL was 1,320 AU, and similarly for TFV above or below 150 ng/mL it was 3,270 AU.

**Figure 1:**
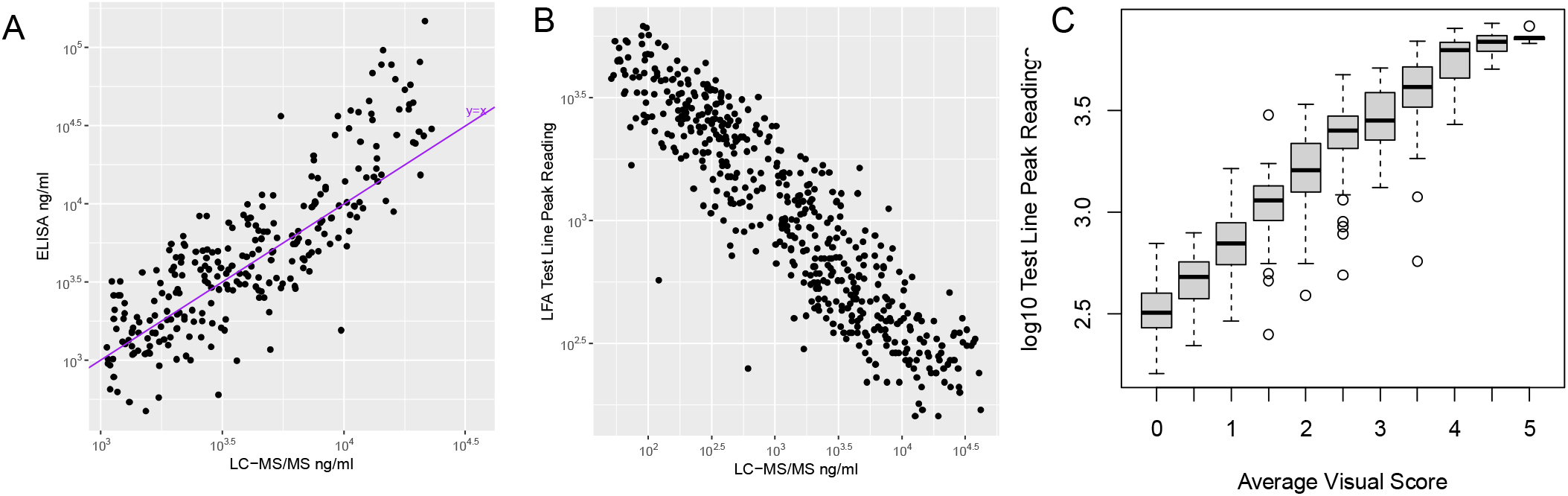
(A) Correlation between ELISA vs LC-MS/MS, for samples with a TFV concentration within the ELISA quantitative of 1,055 ng/mL to 23,000 ng/mL, on log-log axes. The ideal ‘y=x’ line is shown for reference. (B) Correlation between LFA test line (optical reader) with LC-MS/MS, on log-log axis. (C) Correlation of intensity of LFA test line (measured by the optical reader) with visual score, on log10 y-axis.

Regarding visual reads of the LFA, the visual score measured independently by two different people was concordant within 1 grade for 96% of samples. As expected, the average visual score was highly correlated with the measured optical intensity of the test line (**Figure 1C**).

The corrected and uncorrected sensitivities and specificities of all three modalities for both the 1,500 ng/mL and 150 ng/mL thresholds are given in **Table 1**. Altogether, the corrected values were 0 to 4 percentage points higher than the uncorrected values, and do not substantially affect the interpretation of the data. The LFA was found to be no less accurate for differentiating above or below the 150 ng/mL threshold than at the 1,500 ng/mL threshold. At the 1,500 ng/mL threshold, the corrected performance of the scorecard method was likewise statistically similar to that of the reader. Notably, the visual readout of the LFA was not statistically worse for classification at either threshold than ELISA.

**Table 1:** Corrected and uncorrected sensitivity and specificity of the LFA (by average visual score and by optical reader) and the ELISA, at classifying samples above or below two thresholds corresponding to TDF or TAF adherence.

| **1,500 ng/mL TFV Threshold (TDF)** |  |  |  |
| --- | --- | --- | --- |
|  | LFA, Visual (95% CI) | LFA, Reader (95% CI) | ELISA (95% CI) |
| *Uncorrected* |  |  |  |
| Sensitivity | 87% (83% - 91%) | 85% (81% - 89%) | 92% (89% - 95%) |
| Specificity | 92% (88% - 95%) | 96% (92% - 98%) | 90% (86% - 93%) |
| *Corrected\** |  |  |  |
| Sensitivity | 91% (87% - 94%) | 89% (85% - 93%) | 94% (90% - 96%) |
| Specificity | 94% (90% - 97%) | 96% (93% - 98%) | 94% (90% - 96%) |
| **150 ng/mL TFV Threshold (TAF)** |  |  |  |
|  | LFA, Visual (95% CI) | LFA, Reader (95% CI) | ELISA (95% CI) |
| *Uncorrected* |  |  |  |
| Sensitivity | 84% (77% - 90%) | 87% (81% - 92%) | 90% (89% - 94%) |
| Specificity | 89% (85% - 91%) | 95% (92% - 97%) | 83% (79% - 86%) |
| *Corrected<sup>o</sup>* |  |  |  |
| Sensitivity | 87% (80% - 93%) | 90% (84% - 95%) | 92% (86% - 96%) |
| Specificity | 90% (87% - 93%) | 96% (93% - 97%) | 84% (80% - 87%) |
| <sup>a</sup>32 samples within 1275 ng/mL - 1725 ng/mL excluded |  |  |  |
| <sup>o</sup>19 samples within 127.5 ng/mL - 172.5 ng/mL excluded |  |  |  |

## DISCUSSION

To our knowledge, this is the first POC LFA to be useful for monitoring adherence to either TDF or TAF. We found, by using an optical reader, our optimized LFA was just as accurate at the 150 ng/mL TAF threshold as at the 1,500 ng/mL TDF threshold. This result, along with the high correlation between intensity and TFV concentration, also supports the potential feasibility of a future fully-quantitative LFA.

Visual grading using the reference card was also found to be an accurate way to estimate TFV concentration, and may be useful in settings where an optical reader is not available. In the case of TDF, the use of a visual scorecard may be a scalable method to provide additional actionable information about adherence over the previous 2-7 days, which is a longer period than existing ‘yes/no’ tests. More work will be needed to characterize the performance of this approach in diverse clinical settings.

In conclusion, our rapid urine-based LFA is an accurate semiquantitative test of TFV concentration, and can provide real-time information about adherence to either TDF or TAF. When combined with appropriate adherence counseling and interventions, this may be a valuable tool to help reduce HIV transmission and end the global HIV/AIDS pandemic.

## Data Availability

The datasets used and generated in this study are administrated by the corresponding author and may be made freely available by reasonable request.

## Transparency Declarations

None to declare.

## References

1. Mannheimer S, Friedland G, Matts J, Child C, Chesney M. The Consistency of Adherence to Antiretroviral Therapy Predicts Biologic Outcomes for Human Immunodeficiency Virus— Infected Persons in Clinical Trials. Clin Infect Dis. 2002;34(8):1115–1121. doi:10.1086/339074

2. Castillo-Mancilla JR, Morrow M, Coyle RP, et al. Tenofovir Diphosphate in Dried Blood Spots Is Strongly Associated With Viral Suppression in Individuals With Human Immunodeficiency Virus Infections. Clin Infect Dis. 2019;68(8):1335–1342. doi:10.1093/cid/ciy708

3. Drain PK, Bardon AR, Simoni JM, et al. Point-of-care and Near Real-time Testing for Antiretroviral Adherence Monitoring to HIV Treatment and Prevention. Curr HIV/AIDS Rep. 2020;17(5):487–498. doi:10.1007/s11904-020-00512-3

4. Walshe L, Saple D g., Mehta S h., Shah B, Bollinger R c., Gupta A. Physician Estimate of Antiretroviral Adherence in India: Poor Correlation with Patient Self-Report and Viral Load. AIDS Patient Care STDs. 2010;24(3):189–195. doi:10.1089/apc.2009.0208

5. Castillo-Mancilla JR, Haberer JE. Adherence Measurements in HIV: New Advancements in Pharmacologic Methods and Real-Time Monitoring. Curr HIV/AIDS Rep. 2018;15(1):49–59. doi:10.1007/s11904-018-0377-0

6. Hill LM, Golin CE, Pack A, et al. Using Real-Time Adherence Feedback to Enhance Communication About Adherence to Antiretroviral Therapy: Patient and Clinician Perspectives. J Assoc Nurses AIDS Care. 2020;31(1):25–34. doi:10.1097/JNC.0000000000000089

7. WHO | Update of Recommendations on First-and Second-Line Antiretroviral Regimens.; 2019. Accessed October 15, 2019. http://www.who.int/hiv/pub/arv/arv-update-2019-policy/en/

8. Cressey TR, Siriprakaisil O, Kubiak RW, et al. Plasma pharmacokinetics and urinary excretion of tenofovir following cessation in adults with controlled levels of adherence to tenofovir disoproxil fumarate. Int J Infect Dis. 2020;97:365–370. doi:10.1016/j.ijid.2020.06.037

9. Gandhi M, Wang G, King R, et al. Development and validation of the first point-of-care assay to objectively monitor adherence to HIV treatment and prevention in real-time in routine settings: AIDS. 2020;34(2):255–260. doi:10.1097/QAD.0000000000002395

10. Pratt GW, Fan A, Melakeberhan B, Klapperich CM. A competitive lateral flow assay for the detection of tenofovir. Anal Chim Acta. 2018;1017:34–40. doi:10.1016/j.aca.2018.02.039

11. Daughtridge G, Hebel S, Larabee L, et al. Development and Clinical Use Case of a Urine Tenofovir Adherence Test. Presented at the: 2019 HIV Diagnostics Conference; March 28, 2019.

12. Johnson KA, Niu X, Glidden DV, et al. Lower Urine Tenofovir Concentrations among Individuals Taking Tenofovir Alafenamide versus Tenofovir Disoproxil Fumarate: Implications for Point-of-Care Testing. Clin Infect Dis. Published online April 15, 2021.

13. Gandhi M, Bacchetti P, Spinelli M, et al. Brief Report: Validation of a Urine Tenofovir Immunoassay for Adherence Monitoring to PrEP and ART and Establishing the Cutoff for a Point-of-Care Test. Jaids J Acquir Immune Defic Syndr. 2019;81(1):72–77. doi:10.1097/QAI.0000000000001971

14. Ruane PJ, DeJesus E, Berger D, et al. Antiviral Activity, Safety, and Pharmacokinetics/Pharmacodynamics of Tenofovir Alafenamide as 10-Day Monotherapy in HIV-1–Positive Adults. JAIDS J Acquir Immune Defic Syndr. 2013;63(4):449. doi:10.1097/QAI.0b013e3182965d45

15. Panel on Treatment of Pregnant Women with HIV Infection and Prevention of Perinatal Transmission. Recommendations for the Use of Antiretroviral Drugs in Pregnant Women with HIV Infection and Interventions to Reduce Perinatal HIV Transmission in the United States. Accessed April 1, 2021. https://clinicalinfo.hiv.gov/sites/default/files/inline-files/PerinatalGL.pdf

16. Haaland RE, Martin A, Livermont T, et al. Urine Emtricitabine and Tenofovir Concentrations Provide Markers of Recent Antiretroviral Drug Exposure Among HIV-Negative Men Who Have Sex With Men. J Acquir Immune Defic Syndr 1999. 2019;82(3):252–256. doi:10.1097/QAI.0000000000002133

17. Cressey TR, Siriprakaisil O, Klinbuayaem V, et al. A randomized clinical pharmacokinetic trial of Tenofovir in blood, plasma and urine in adults with perfect, moderate and low PrEP adherence: the TARGET study. BMC Infect Dis. 2017;17(1):496. doi:10.1186/s12879-017-2593-4

18. Drain PK, Kubiak RW, Siriprakaisil O, et al. Urine Tenofovir Concentrations Correlate With Plasma and Relate to Tenofovir Disoproxil Fumarate Adherence: A Randomized, Directly Observed Pharmacokinetic Trial (TARGET Study). Clin Infect Dis. Published online 2019. doi:10.1093/cid/ciz645

19. Sevenler D, Bardon A, Fernandez Suarez M, et al. Immunoassay for HIV Drug Metabolites Tenofovir and Tenofovir Diphosphate. ACS Infect Dis. 2020;6(7):1635–1642. doi:10.1021/acsinfecdis.0c00010

20. 2020 HIV Market Report. Clinton Health Access Initiative; 2020. Accessed April 2, 2021. https://www.clintonhealthaccess.org/hiv-mid-year-market-memo-2020/

